# Trained dogs can detect the odour of Parkinson’s Disease

**DOI:** 10.1101/2023.11.01.23296924

**Authors:** Nicola Rooney, Drupad K Trivedi, Eleanor Sinclair, Caitlin Walton Doyle, Monty Silverdale, Perdita Barran, Tilo Kunath, Steve Morant, Mark Somerville, Jayde Smith, Julie Jones-Diette, Jenny Corish, Joy Milne, Claire Guest

**Author notes:** Correspondence address: Animal Behaviour and Welfare Group, Bristol Veterinary School, Langford, UK. BS40 5DU.

## Abstract

A definitive diagnostic test for PD remains elusive, so identification of potential biomarkers may shed light on methods for diagnosis and facilitate early intervention. Excess sebum secretion and skin pathology are recognised symptoms of early PD. It is likely these result in a unique signature of volatile organic compounds that could be used to identify early stages of disease. Numerous medical conditions produce distinctive odours, and dogs have been trained to detect many of these. A single previous study, suggested that dogs can also be trained to detect Parkinson’s Disease. In this study, two dogs were trained to distinguish sebum swabs obtained from drug naïve, and medicated Parkinson’s patients from swabs from control participants. After 38-53 weeks of training on 205 samples (90 target and 115 control), the dogs were tested in a double-blind trial using 60 control and 40 target samples from drug-naïve patients. The dogs both showed high sensitivity (proportion of target samples found 70% and 80%) and specificity (proportion of control samples not alerted to 90% and 98%) of alerting response. This trial supports previous findings that dogs can be trained to reliably detect the odour of PD. We suggest there is a potential for dogs to achieve even higher accuracy with increased exposure and refined training methods and to detect early-stage PD, even prior to diagnosis, as well as hard to diagnose PD cases. Further exploration of the factors which affect dogs’ sensitivity and specificity and sample features which affect accuracy of discrimination are now required.

## Introduction

Parkinson’s disease (PD) is a neurological disease that causes progressive motor symptoms including tremor, stiffness and slowness of movement, as well as non-motor symptoms including mood problems, cognitive issues, sleep disturbance and pain. Neurodegeneration, predominantly in the central nervous system is the cause to these widespread symptoms. At Braak stage 3, the Substantia Nigra becomes affected, leading to a reduction in dopamine production in the brain, a chemical that is vital for the control of movement^**Error! Reference source not found**.^. As progression of the disease can be slow, often with only very minor symptoms initially, it is difficult to definitively diagnose PD particularly in the early stages. Early motor and non-motor symptoms, may result in people being diagnosed with a range of other conditions including rheumatological, neurological, sleep and mood disorders^2^. Even as the disease progresses, it can be difficult to differentiate from other Parkinsonian disorders^3^. Early and accurate diagnosis is vital, to allow patients to benefit from pharmacological interventions, such as a levodopa/carbidopa, which may be more effective when administered early^4^. Non-pharmacologic interventions, such as exercise, are also easier for the patient to perform before declines in locomotor skills and movement occur^4^.

Around 1 in 500 people in the UK are affected by PD, a prevalence of approximately 127,000 in 2017^5^. Gumber et al. (2017) suggests an economic burden to the NHS of £2,118 per person with Parkinson’s (PwP), hence the total UK annual cost could be in excess of £268 million per year ^5^. With an aging population and rising demand for health services, this will create a significant challenge for healthcare providers. Research is needed to support treatment decisions, direct prevention strategies, raise awareness, and avoid unnecessary and expensive repeat admissions between home and hospital.

Studies that aid our understanding of early symptoms could help develop early diagnosis are paramount. Much research has attempted to identify new biomarkers^6^, particularly those that may predict whether someone will develop PD in the future.^7^ One early non-motor symptom, is seborrheic dermatitis: excessive sebum (waxy or oily secretion) which is excreted from the sebaceous glands at higher quantities in patients with PD ^8, 9^. Sebum has been noted to have a very distinctive odour identified in repeated blinded studies with human super-smeller Joy Milne, who has hyperosmia. This could potentially be used as an indicator for the disease ^10^ and is associated with lipid dysregulation consistent with many PD studies^11^.

Many medical conditions have distinctive odours ^12,13^ including cancer ^14^ and schizophrenia ^15,16^. Health care workers have reportedly used scent to triage illness, and in studies as early as 1960, when it was shown that sweat from schizophrenic and non-schizophrenic patients could be distinguished by trained rats using odour alone^17^. There is a growing body of evidence that disease-specific volatile organic compounds (VOCs) could be used for the early detection of disease before more obvious clinical signs or symptoms become apparent^18^. The mechanism for the release of volatiles is not well studied and varies with disease. However, small amounts of volatile organic compounds are released through metabolic processes and can be identified in the secretions of sweat, mucus, sebum, urine, blood and breath ^19^.

The olfactory capability of dogs to detect diseases is well-established ^20^. For over twenty years, the UK charity Medical Detection Dogs (MDD) has trained dogs to detect human diseases ranging from prostate cancer ^21^ to malaria ^22^ and Covid-19 ^e.g. 23^. MDD has also trained assistance dogs to alert to the onset of life-threatening episodes for people living with chronic diseases such a Type 1 diabetes ^24-25^. It therefore seems extremely plausible that dogs could be trained to alert to the odour of PD.

A recent prospective, diagnostic case-control study was conducted in China ^28^. Three dogs trained in a laboratory were tested on sebum samples from PD medicated and drug naïve patients, and matched controls. The diagnostic criterion applied was that at least two of the three dogs alerted to the sample and, using this criteria, the study saw sensitivity and specificity rates of 91% and 95% to medicated and 89% and 86% to drug naïve samples, respectively. The authors concluded that trained dogs could be a useful, quick non-invasive and cost-effective method to identify patients with PD during community screening and health check-ups as well as within neurological practice, but provide limited information on the dog’s training and testing.

In the current study, we further tested the capacity of trained dogs to discriminate the odour of Parkinson’s disease from controls using sebum swabs. Since the dog is effectively the measuring instrument, it is important to provide full details of its training, calibration and testing which we describe fully. To simulate real operations, unlike Gao et al ^28^, we did not use precisely matched control samples. However, since it is vital to minimise all possible confounding factors ^e.g. 29^ we trained the dogs using samples taken from drug naïve (DN) patients shortly after diagnosis and a control group ranging from healthy individuals to patients with other neurological conditions. We started with ten potential dogs, and based on their performance during initial training, we selected two dogs, which were trained by a professional dog trainer (MS) using an established protocol ^21^ and then tested during double-blind trials.

## Materials and Methods

### Ethical Review

Ethical approval for this project (IRAS project ID 191917) was obtained by the NHS Health Research Authority (REC reference: 15/SW/0354).

### Participant Recruitment

Parkinson’s disease patient and control participant samples were collected during a nationwide recruitment at 25 different NHS clinics as described in Trivedi et al. ^10^. The participants for the current study were selected at random from those collected for that study, in order to match phenotypes as closely as possible for both training and testing. Those used for testing were all drug naïve, whilst medicated patients were also used in the latter stages of training. The control samples were collected from people accompanying PwP in clinics at the same sites and additional control samples came from volunteers attending the Medical Detection Dog centre.

### Sample collection

All participants were asked to avoid showering, use of cosmetics and fragrances for 24 hours prior to sample collection to minimise contamination from washing cosmetics. Sampling followed the same standard operating procedure for disease and control study volunteers. Sebum was sampled from volunteers by wiping a square piece of medical gauze along the hairline, down the nape of the neck and across the shoulder, this was performed by a clinician for samples taken in NHS clinics and by the volunteer themselves for other controls. The upper trunk was selected as it is a sebum rich location and during pilot method development studies, this area was indicated by human super smeller to have highest odour linked to PD^19^ .The gauze samples were bagged individually and sealed in an airtight, minimal odour, plastic bag which was labelled with the participants’ anonymised code. Samples were shipped to a central Manchester facility and stored at -80°C within 24 hours of arrival.

From here the samples were transported on dry ice to the dog training centre accompanied by a recruitment cohort code, participant sex, age, BMI, ethnicity, medical conditions, current medications, drinking and smoking behaviour (obtained from a questionnaire that participants filled in at the time of sampling) and whether they were a control or patient. For patients, further information was provided on date of diagnosis, and medications they were taking for PD. All samples were stored at -80 °C.

In total, 130 samples were collected from PD patients and 175 from control and these were randomly divided into 105 training and 100 testing samples. All samples were cut in half ahead of storage. Duplicate samples were either both used within the training samples, or one was used as a training sample, and the other as a “filler” control during testing, to enable lines of four samples to be presented but the dogs’ response to any “familiar” samples during testing was not analysed. The training samples were labelled as positive or control, but the test samples were given only a code, so testing was totally blind.

### Canine Disease Detection Training

After initial screening of ten dogs, five were deemed to have potential aptitude and hence commenced initial PD training.

A training protocol was used which increased in difficulty over time. In Phase 1, the target scent was introduced to the dog, and the dog rewarded for showing a behavioural change in response. Individual dogs naturally show different behaviours, and these were rewarded by their trainers, so each dog gave a consistent behaviour e.g., sit or stand and stare, thereafter terms an indication. Correct responses were rewarded with food or ball-play, in conjunction with the use of an audible clicker (used as a secondary reward).

Initial training used search and find games, then dogs were trained to walk along a line of four stainless steel stands, one metre apart, each with an arm holding a 40 ml glass jar containing a swab sample. A stainless-steel grill was placed over the mouth of each jar to prevent the dog touching the sample. After each line was searched, the plates were cleaned in an ultrasonic cleaner with an enzymatic solution (Reprozyme TM, NVS Stoke-on-Trent UK), the plates were rinsed thoroughly using a commercial glass washer (minimum 85 °C) then left to air dry to avoid any contamination. The jars were cleaned in the same way but additionally were placed in an oven at 180-C for 20 minutes.

The dog was trained to sniff each of the four stands in order. Their behaviour at each stand was recorded on a database as IND (indicated), HES (hesitated), INT (interest, but weaker than HES), NI (no interest) or NS (not searched, usually because a sample earlier in the line was indicated). Initially dogs were presented with one positive sample and three control samples during each run. As training progressed, lines with four controls were also presented. Dogs were rewarded for both indicating positive target samples and for searching a negative line without an indication. Once dogs were reliably indicating positive target samples against control samples taken from participants with non-neurological conditions, we added control samples taken from patients with diseases with similar symptoms to Parkinson’s disease, including depression, anxiety, bi-polar disorder, epilepsy, stroke, hypothyroidism and migraine. Later training also included medicated (as well as drug naïve) target samples to ensure dogs were discriminating entirely based on PD status.

As training progressed, three of the dogs failed to learn the task, and hence were rejected from training. Only the data from the two remaining dogs is presented here. They were a two-year old male Golden Retriever (Dog 1) and a three-year old male Labrador x Golden Retriever (Dog 2).

In total, we used 205 samples during training; 90 targets and 115 controls. Dog 1 took part in 72 training sessions over 38 weeks and encountered control samples on 1683 occasions and target samples on 597 occasions. Dog 2 took part in 83 sessions over 53 weeks and encountered control samples on 1759 occasions and target samples on 582 occasions. Each training session lasted an average of 21 minutes and a maximum of 69 minutes. On three occasions during training, both dogs indicated in response the same control sample, and in these cases the sample was withdrawn from the training process.

### Canine Disease Detection Testing

The two dogs that were deemed to have learnt the task adequately, proceeded to a double-blind randomised controlled trial and were presented with 100 previously unseen samples, of which 40 were from individuals with confirmed disease (positive for PD). Due to the scarcity of controls, we used an additional 100 duplicates of control samples seen by the dogs during training. These were used as “fillers” to facilitate presenting lines of four samples, but the dogs’ responses to these familiar samples were not included in any analyses.

The samples were presented in the same way as during training in a four-stand alignment. In each “run” there was either one or no positive samples. Both the trainer/handler and the experimenter were blind to the presence and position of any targets. Ten control and five positive samples from the training set were used for calibration runs, to confirm the dog was focused on the scent. Calibration runs were requested at any time by the trainer, were unblinded and their results were not included in the analysis.

The regular trainer guided the dog to search each line of samples. Dogs were permitted to make a maximum of four passes over the line. Once the handler was confident of the dog’s choice, a decision was communicated to the experimenter. For each the dogs’ behaviour was coded as it was during training (IND, HES, INT, NI or NS), with HES, INT and NI all being treated as a negative response for analysis.

Each line of samples was presented in reverse order on a second occasion, so that samples for which no decision was made in the first run re-presented. Any samples that were still not seen after this run were collected together in new lines, until a decision had been made for all samples. This process resulted in some samples being encountered more than once by the same dog, however we only analysed their response on first encounter.

### Data Management and Analysis

A blinded experimenter entered the responses to each sample into the database. All trial samples had previously been given a “blind ID” which was read by the data collection software to determine whether responses were correct. The software revealed whether each response was correct, and if so, the trainer was asked to reward their dog. If any sample was incorrect, no reward was given. The database was password protected and only the trial statistician (SM) knew both passwords prior to trial completion.

We used the first encounter with each of the 100 novel samples to calculate sensitivity (proportion of target samples found) and specificity (proportion of control samples that were not alerted to) for each dog. We also tested whether there was any association between the errors made by each dog, examining target and control samples separately using Fishers Exact tests.

## Results

### Training

The performance of two dogs during training was deemed satisfactory. In its last 10 training sessions Dog 1 responded correctly to 91% of encounters and Dog 2 responded correctly to 90% of encounters.

### Testing

Both dogs demonstrated an ability to discriminate between the PD positive and control samples they had not encountered before (*P*<0.0001), with sensitivities of 70.0% and 80.0% and specificities of 90.0% and 98.3% (Table 1).

**Table 1.** Sensitivity and specificity of two dogs in double-blind trials.

|  | Samples |  | Indications |  | Sensitivity and specificity with confidence intervals (%) |
| --- | --- | --- | --- | --- | --- |
|  | Type | *n* | Number indicated | % indicated (as PD) |  |
| **Dog 1** | Control | 60 | 6 | 10.0 | Specificity 90.0 (79.5, 96.2) |
|  | Positive | 40 | 28 | 70.0 | Sensitivity 70.0 (53.5, 83.4) |
|  |  |  | *p*<0.0001 |  |  |
| **Dog 2** | Control | 60 | 1 | 1.7 | Specificity 98.3 (91.1, 100.0) |
|  | Positive | 40 | 32 | 80.0 | Sensitivity 80.0 (64.4, 91.0) |
|  |  |  | *p*<0.0001 |  |  |
| *p*-values for Fisher’s exact tests for testing the difference in indication rates between target and control samples. |  |  |  |  |  |

**Table 2.**
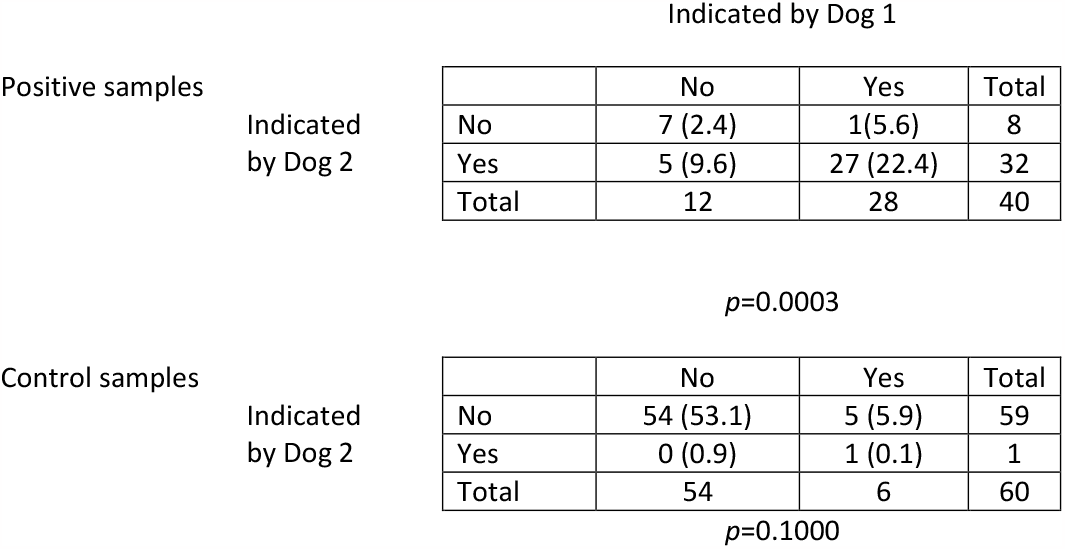
Agreement between responses of the two dogs during double blind testing p-values for Fisher’s exact tests. Numbers in brackets are the expected numbers if there were no correlation between the dogs’ responses.

|  |  | Indicated by Dog 1 |  |  |  |
| --- | --- | --- | --- | --- | --- |
| Positive samples | Indicated<br>by Dog 2 |  | No | Yes | Total |
|  |  | No | 7 (2.4) | 1(5.6) | 8 |
|  |  | Yes | 5 (9.6) | 27 (22.4) | 32 |
|  |  | Total | 12 | 28 | 40 |
| $p=0.0003$ |  |  |  |  |  |
| Control samples | Indicated<br>by Dog 2 |  | No | Yes | Total |
|  |  | No | 54 (53.1) | 5 (5.9) | 59 |
|  |  | Yes | 0 (0.9) | 1 (0.1) | 1 |
|  |  | Total | 54 | 6 | 60 |

$p=0.1000$

Agreement between the two dogs was higher than expected by chance for positive samples: 7 were missed by both dogs (expected 2.4) and 27 were indicated by both dogs (expected 22.4). Agreement between the dogs for control samples was close to expectation.

## Discussion

This study confirms the previous findings of Gao et al ^28^ that dogs can be trained to discriminate between Parkinson’s patient and control sebum swab samples. Although only two dogs from the initial ten trialled were deemed suitable to progress to full training, the subsequent performance of these during training and double-blind testing demonstrates that it is possible for dogs of the correct aptitude to learn the task.

The two dogs achieved high sensitivity and specificity, when trained using ex-situ sebum samples, showing there is an olfactory signature which is different in patients with Parkinson’s Disease. Since increased sebum secretion has also been noticed before motor signs of PD ^30, 31^ this suggests that detection of biomarkers in sebum could lead to early diagnosis and intervention.

Agreement between dogs was higher than would be expected by chance, suggesting that some samples were easier to identify than others. There were several samples to which both dogs incorrectly indicated. Although initial inspection of sample details suggested no obvious commonalities between these samples, further research is required to understand which features of the sample are responsible for these differences.

Sensitivity levels of 70 and 80% are high; well above chance and comparable to levels seen for example in malaria detection ^22^, and considerably higher than those obtained when using dogs to detect some other medical conditions (e.g., 41% bladder cancer ^21^). The only other study examining detection of PD by dogs, obtained higher sensitivity (91%), but similar specificity (95%) as seen here (98% and 90%).

There are multiple plausible reasons for the difference in sensitivity between the two studies; Gao et al ^28^ used consensus responses of two out of three dogs as the detection criterion ^28^ whilst our analysis took each of two dog’s responses individually. Gao’s testing protocol used PD samples and six perfectly matched controls, whilst ours included controls that were not precisely matched which may make the task harder for the dogs, but we deem this to be more authentic. Gao et al also presented a known PD target to the dogs ahead of each testing run so dogs were able to “match to source”, which may improve their detection but may not be practicable in an operational setting.

The value of this procedure is yet to be determined and is an important area of future study. Gao’s dogs had been trained on PD over a period of two years, whilst ours were trained for less than a year, although the number of samples encountered during training was not described by the Gao at al., so cannot be compared There were several other details of the dog testing methods which were not described by Gao et al, such as the period over which testing occurs, or a description of the “tanks” in which samples were presented, which precluding further comparisons. However, the studies together suggest that refinements to training and sample presentation in the future could lead to further increases in detection accuracy.

In our study, whilst the specificity of Dog 2 was 98% reflecting very few false alerts, and similarly the dog showed the highest sensitivity (80%), Dog 1 showed slightly lower specificity (90%) and a sensitivity of only 70%. These differences may reflect personality differences between the two dogs, with Dog 1 showing a more liberal approach to responding; a higher tendency for false alarms thus minimising false negatives to increase the chance of a hit ^32^. Or it may reflect subtle differences in their training, leading Dog 2 to be more precise in its responses. Further analysis using the data stored during training, will enable understanding of the extent to which these individual differences develop or are consistent from early training.

In conclusion, the sensitivity and specificity of dogs trained to find PD positive samples is encouraging and supports previous finding that further research has the potential to help develop methods for the early diagnosis of PD, by dogs or by other technology. This may in turn lead to opportunities for earlier intervention than is possible at present and be especially beneficial for hard to diagnose cases of PD. Longitudinal studies of dogs’ capacity to alert prior to formal diagnosis are now required.

## Data Availability

The data supporting the findings of this study are openly available at the University of Bristol data repository, data.bris, at https://doi.org/10.5523/bris.2ffiar75hr0b82ottk9axxh44i

## Acknowledgements including source of funding

We thank Michael J Fox Foundation and Parkinson’s UK for funding this study. We also thank the nationwide NIHR recruitment centres for their support and would like to thank the patient’s carers and participants for providing the samples. We are very grateful to all the participants who took part in this study.

## Author contributions statement

CG, PB, MS and TK conceived the study. CG, JC, KW designed the experiments. CG, and PB supervised the study. DT., ES., CWD, JM., and PB performed or contributed to the sample collection and preparation. MS performed the dog sample preparation, dog training and data collection, with assistance from JS. SM performed statistical analysis. NJR wrote the manuscript, with input from JJD. All authors commented and contributed to the final version of manuscript.

